# Efficacy and safety of integrated Chinese and western medicine for cancer therapy-induced thrombocytopenia: A meta-analysis

**DOI:** 10.1101/2025.10.30.25338849

**Authors:** Yanying Zhang, Fang Liu, Xuejiao Ma, Li-qun Jia

## Abstract

**Objective:** To evaluate the efficacy and safety of Traditional Chinese medicine (TC M) combined with Western medicine (WM) for cancer therapy-induced thrombocytopenia (CTIT).

**Methods:** A comprehensive literature search was performed across multiple databases, including PubMed, Embase, Cochrane Library, China biology medicine, China national knowledge infrastructure, Wanfang, and VIP, from inception to August 2025. Randomized controlled trials (RCTs) examining the effects of integrated TCM and WM on CTIT were included. The primary outcomes were significant response rate and overall response rate, while secondary outcomes included nadir platelet count, duration of thrombocytopenia, and adverse events. The methodological quality of the included RCTs was assessed using the Cochrane Risk of Bias Tool (version 5.1.0).

**Results:** A total of 33 studies involving 2,443 patients were included, comprising 1,204 patients in the experimental group and 1,186 in the control group. All studies were conducted in China and published between 2004 and 2024. Compared with WM alone, integrated TCM and WM significantly improved overall response r ate (risk ratio [RR] = 1.40, 95% confidence interval [CI]: 1.14-1.73), increased plate let counts (RR = 13.59, 95% CI: 11.01-16.17), shortened the duration of thrombocytopenia (MD = -3.46, 95% CI: -4.30 to -2.63), and reduced the incidence of adverse events (RR = 2.34, 95% CI: 1.52-3.6).

**Conclusions:** In conclusion, combined TCM and WM demonstrates significant benefits in alleviating CTIT, elevating platelet counts, reducing thrombocytopenia duration, and minimizing side effects associated wit h thrombopoietic therapies compared to WM alone.

## 1. Introduction

Cancer treatment-induced thrombocytopenia (CTIT) is a prevalent adverse effect of anti-tumor therapies, with its incidence influenced by factors such as tumor type, treatment regimen, and chemotherapy duration. Studies have reported an incidence rate as high as 21.8% in patients undergoing anti-cancer treatment^[1]^. CTIT not only elevates the risk of bleeding, prolongs hospital stays, and increases healthcare costs, but it can also be life-threatening. Current Western medical interventions for thrombocytopenia primarily include recombinant human interleukin-11 (rhIL-11) and recombinant human thrombopoietin (rhTPO). In cases of insufficient response, thrombopoietin receptor agonists may be considered, and platelet transfusions are administered when necessary to prevent severe hemorrhage. Immunotherapeutic approaches, such as corticosteroids and intravenous immunoglobulin, are also utilized in the treatment of immune thrombocytopenia ^[2]^. Although these interventions effectively elevate platelet counts, they have significant limitations: rhIL-11 is associated with cardiotoxicity risks ^[3]^, rhTPO requires liver function monitoring, and repeated transfusions may lead to platelet alloimmunization. Furthermore, the high costs of these treatments impose a considerable eco nomic burden on patients. Thus, identifying novel therapeutic strategies for managing CTIT is of paramount importance.

Despite the widespread use of integrated Traditional Chinese and Western Medicine (TCM-WM) in the management of CTIT ^[4]^, the current evidence supporting its efficacy and safety remains limited. This limitation is underscored by the latest expert consensus, which explicitly highlights the low quality of evidence for TCM interventions, primarily due to the lack of high-quality randomized controlled trials (RCTs). Consequently, there is an urgent need for more rigorous and synthesized evidence to guide clinical decision-making. The present systematic review and meta-analysis, therefore, aims to consolidate existing clinical trial data to evaluate the therapeutic value of TCM-WM integration in treating CTIT, with the ultimate goal of providing higher-level evidence to address this gap in knowledge. This review was registered with PROSPERO (Registration number: CRD420251083197) and conducted in accordance wit h the PRISMA guidelines ^[5]^.

## 2. Methods

### 2.1. Search strategy

Seven databases, including PubMed, Embase, Cochrane Library, China Biology Medicine (CBM), China National Knowledge Infrastructure (CNKI), Wanfang, and VI P, were systematically searched from their inception through August 2025. All randomized controlled trials (RCTs) involving patients with cancer treatment-induced thrombocytopenia (CTIT) were considered for inclusion. The search terms included “Thrombocytopenia,” “Thrombocytopenias,” “Thrombopenia,” and “Traditional Chinese Medicine,” among others. A detailed search strategy is presented in Table S1. Article screening was performed independently by two authors, with discrepancies resolved by a third author.

### 2.2. Inclusion criteria and exclusion criteria

The inclusion criteria were as follows: (1) Participants: Adults aged 18-75 years, or older, with thrombocytopenia induced by cancer therapy; (2) Interventions: Integration of Traditional Chinese Medicine (TCM) and Western Medicine (WM) therapies; (3) Comparisons: Western Medicine therapies, with or without placebo; (4) Outcomes: The primary outcomes were the significant response rate and overall response rate, as defined by the National Cancer Institute grading criteria for acute and subacute toxicity of anticancer agents (Table S2). Secondary outcomes included nadir platelet count, duration of thrombocytopenia, and adverse events; (5) Study design: Randomized controlled trials (RCTs) with two arms, consisting of an intervention group and a control group.

Studies were excluded if they used inappropriate outcome measures for overall response rate or were not published in English or Chinese.

### 2.3. Data extraction and quality assessment

Two investigators independently screened all eligible studies to extract the following data: authors, year of publication, sample size, patient age, interventions in the experimental and control groups, treatment duration, study locations, and outcomes, including significant response rate, overall response rate, nadir platelet count, duration of thrombocytopenia, and adverse events. The methodological quality of the studies was assessed based on key factors, including randomization, allocation concealment, blinding of outcome assessment, completeness of outcome data, selective reporting, and other potential sources of bias. The risk of bias was evaluated following the guidelines outlined in the Cochrane Handbook for Systematic Reviews of Interventions (version 5.1.0).

### 2.4. Data analysis

Data analysis was performed using the statistical software STATA SE 16.0. Dichotomous data were presented as risk ratios (RR), and continuous outcomes as mean differences (MD), both accompanied by 95% confidence intervals (CIs). Statistical heterogeneity was evaluated using the I² statistic, with I² < 25% indicating low heterogeneity, 25% ≤ I² ≤ 50% indicating moderate heterogeneity, and I² > 50% indicating high heterogeneity. An I² > 50% or a P-value < 0.1 suggested the presence of significant statistical heterogeneity. A random-effects model was employed for meta-analysis in the presence of substantial heterogeneity, while a fixed-effects model was used when heterogeneity was minimal.

## 3. Results

### 3.1. Search results

A total of 490 potentially relevant articles were identified through title and abstract screening after removing 34 duplicates. Following initial screening, 409 articles were excluded, including 102 reviews or basic/mechanistic studies and 327 studies not related to CTIT. The full texts of the remaining 81 articles were assessed, and 48 were further excluded: 13 did not meet the outcome criteria, 10 lacked reported outcomes, and 19 were not RCTs. Ultimately, 33 studies ^[6–38]^ involving a total of 2,443 participants met the inclusion criteria. The study selection process is illustrated in the flow chart (Fig.1).

**Fig. 1.**
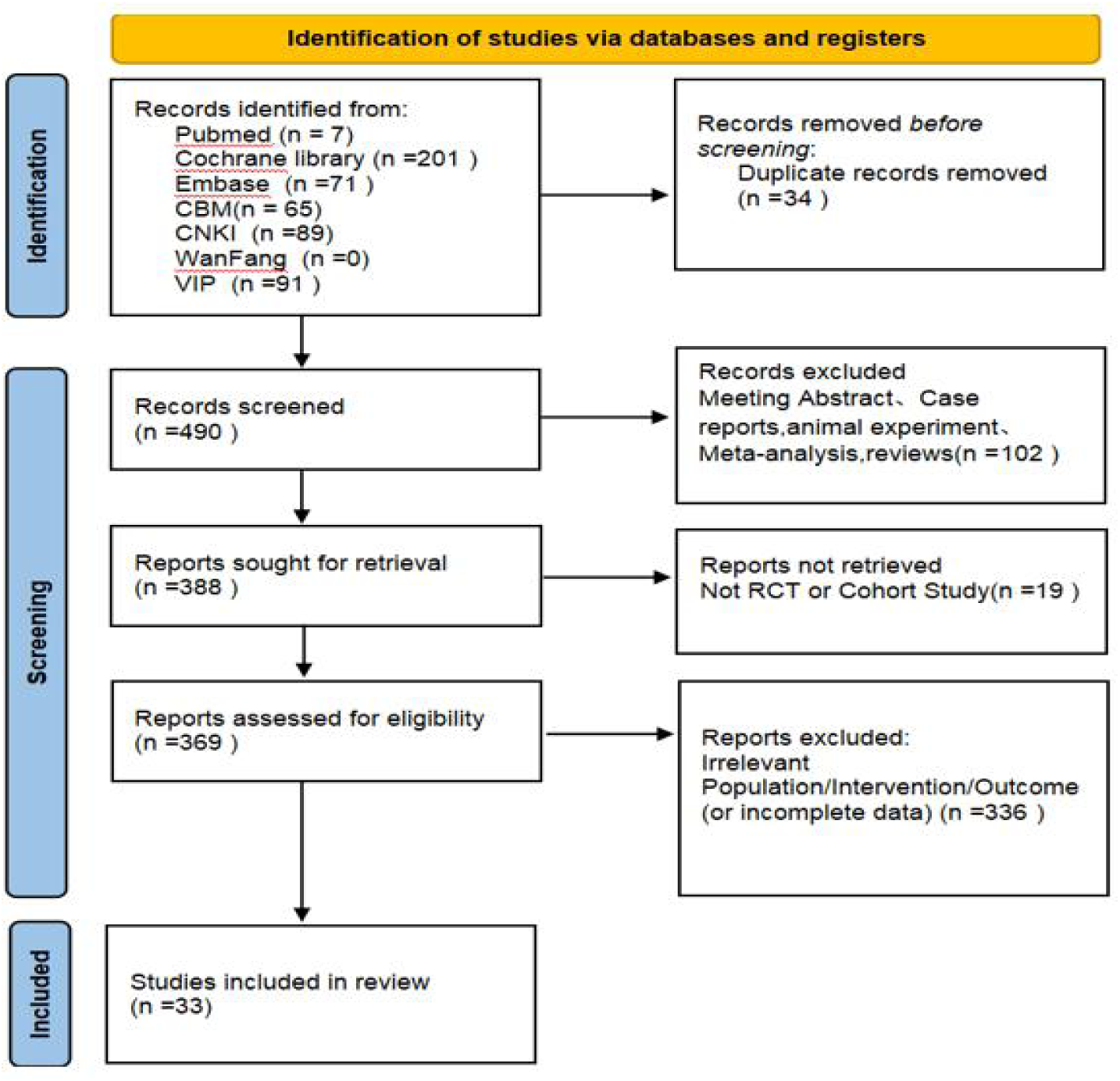
PRISMA Flow diagram.

### 3.2. Study characteristics

A total of 2,443 patients were included across 33 studies, with 1,204 patients in the experimental group and 1,186 in the control group. All studies were conducted in China and published between 2004 and 2024. Nineteen studies designated significant response rate as the primary outcome, while 20 studies identified overall response rate as the primary outcome. Nadir platelet count following treatment was reported in 21 studies, and the duration of thrombocytopenia was documented in 18 studies. A summary of the study characteristics is provided in Table 1.

**Table 1.**
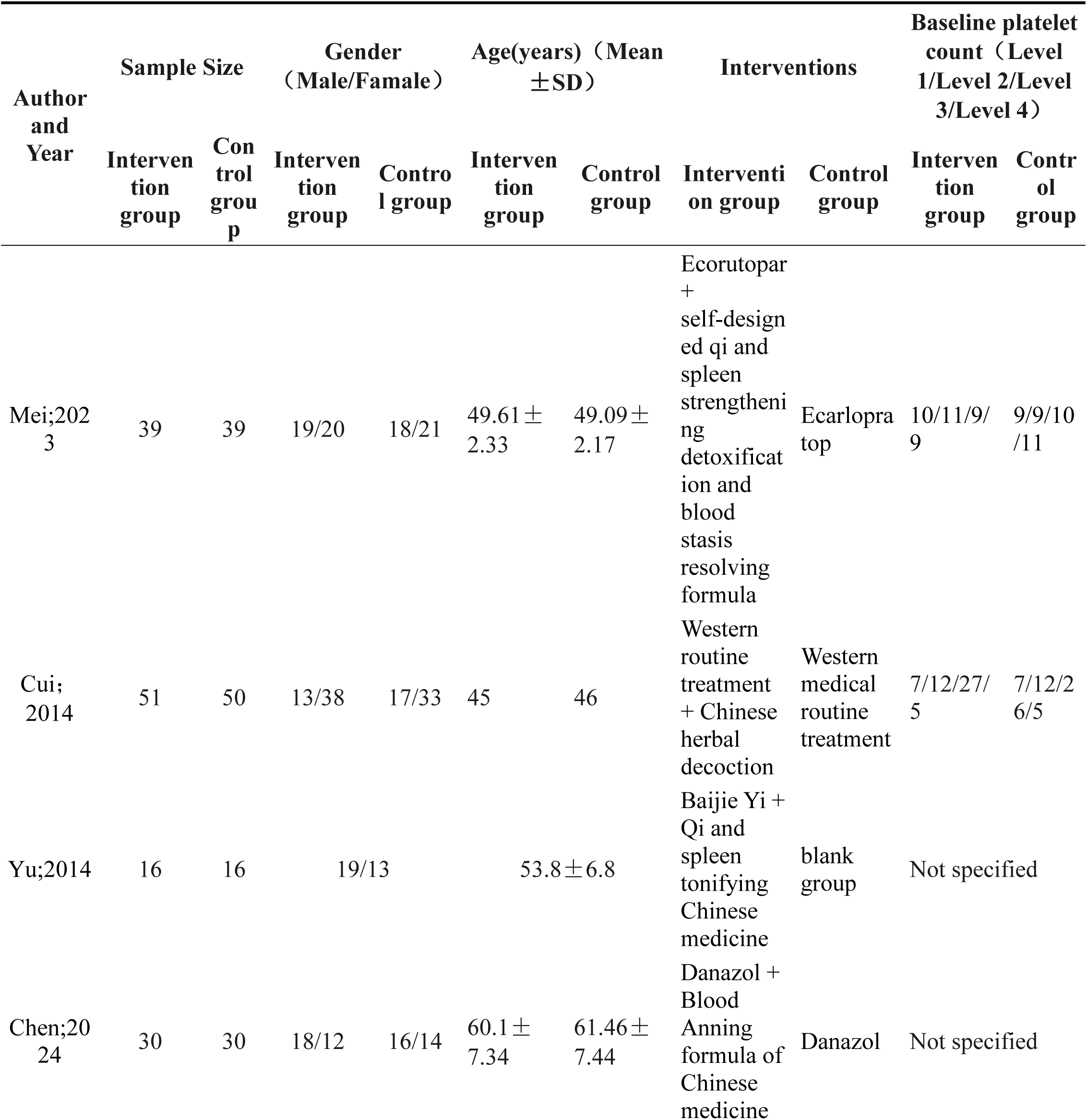
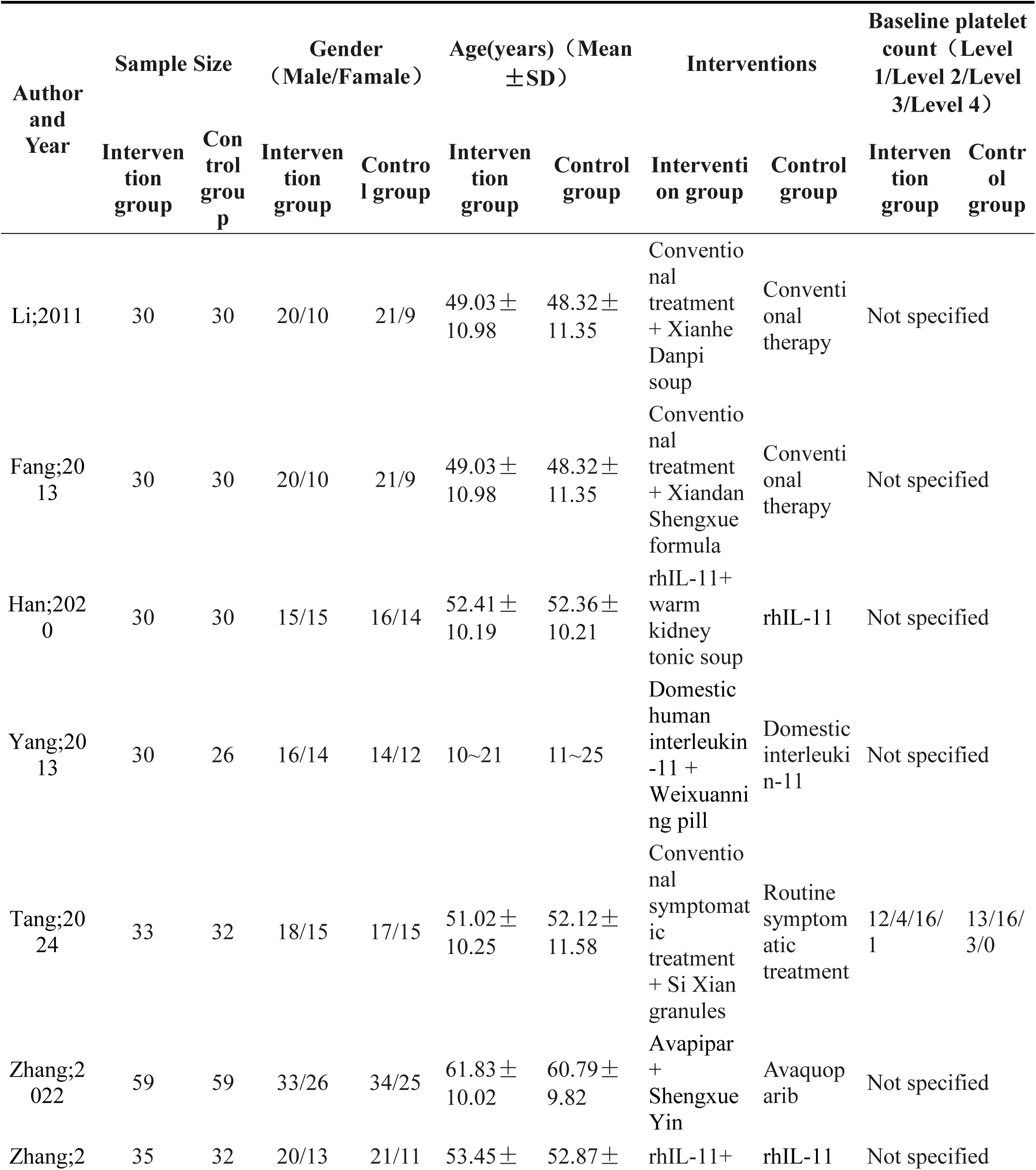
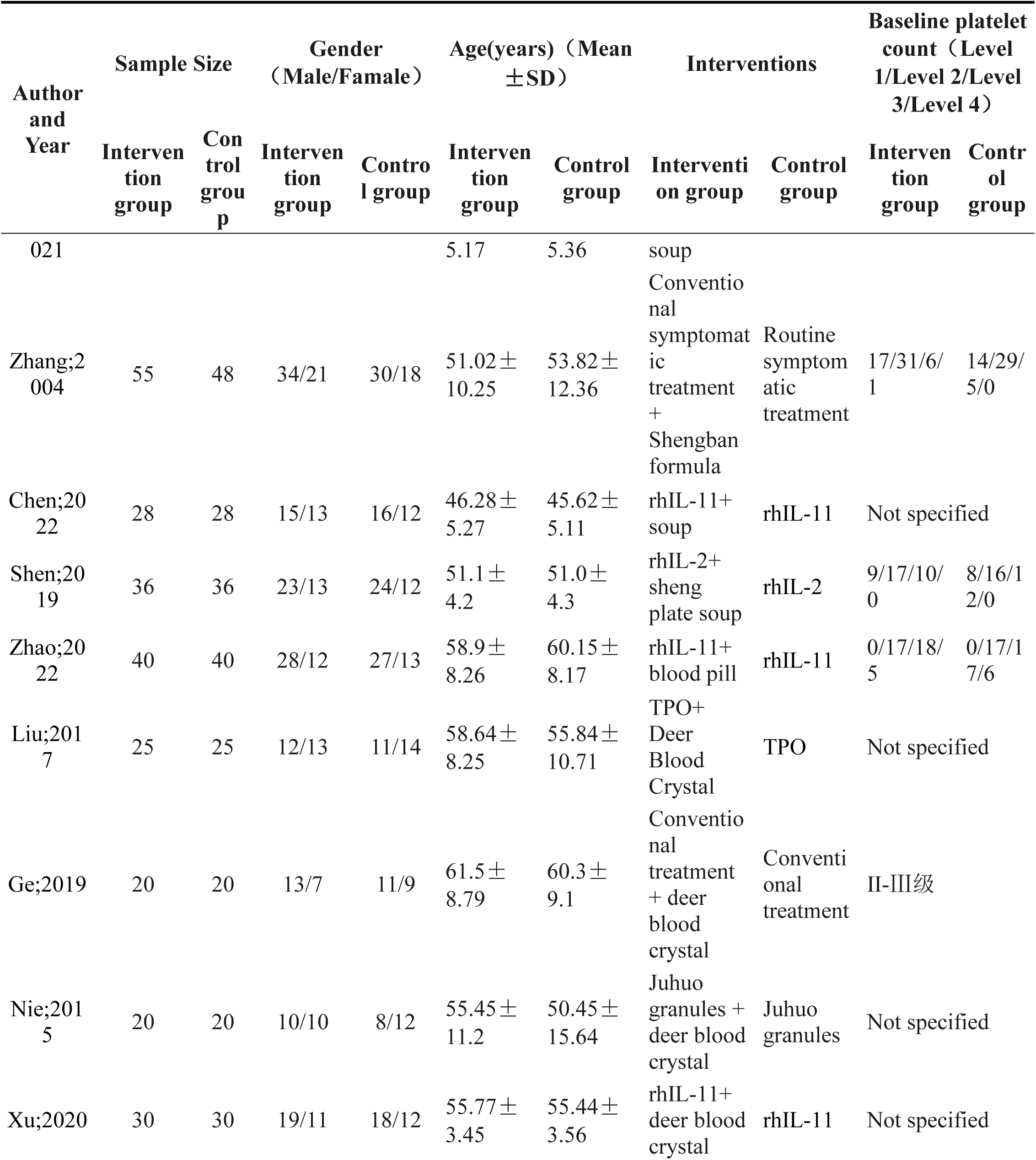
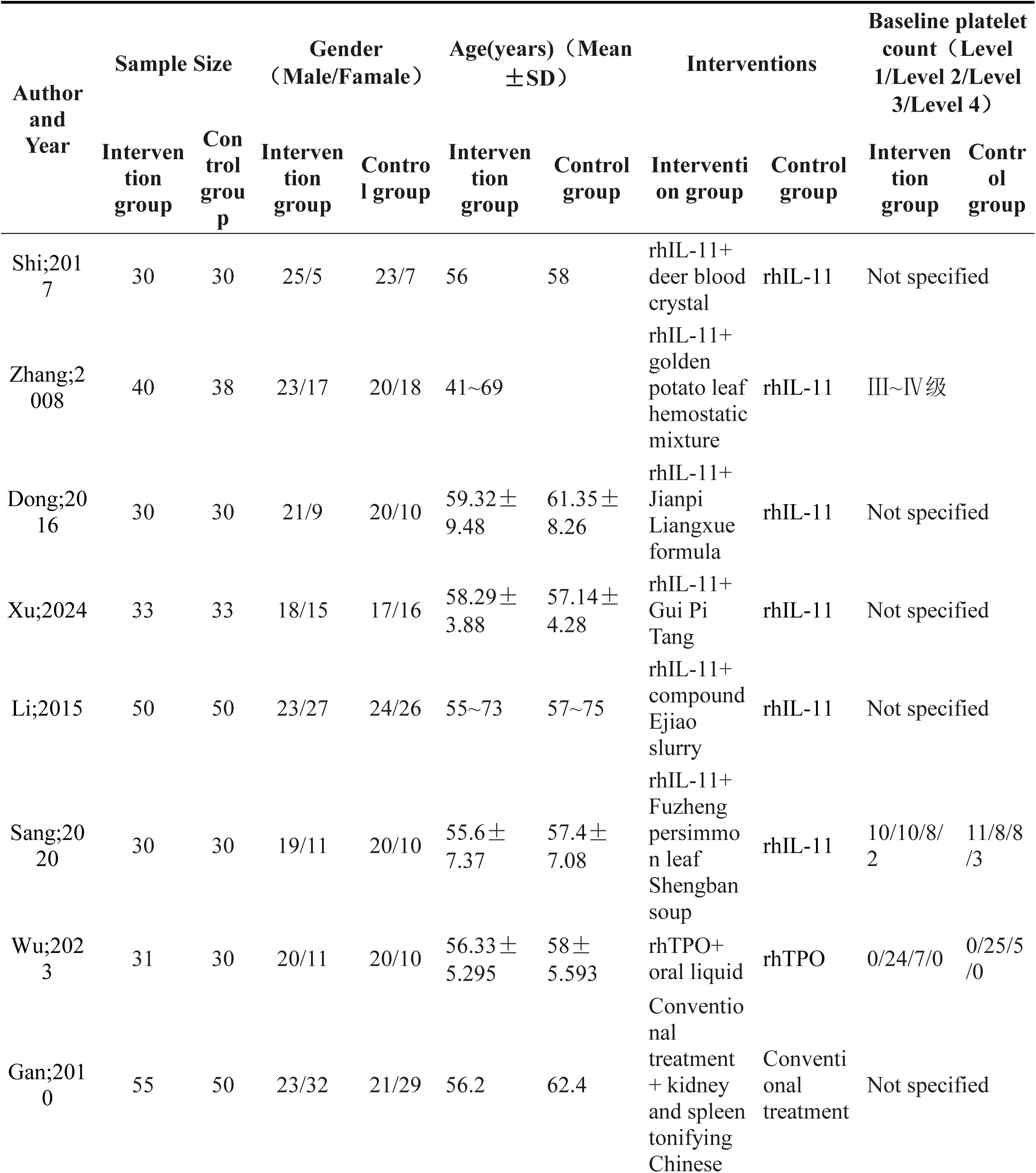
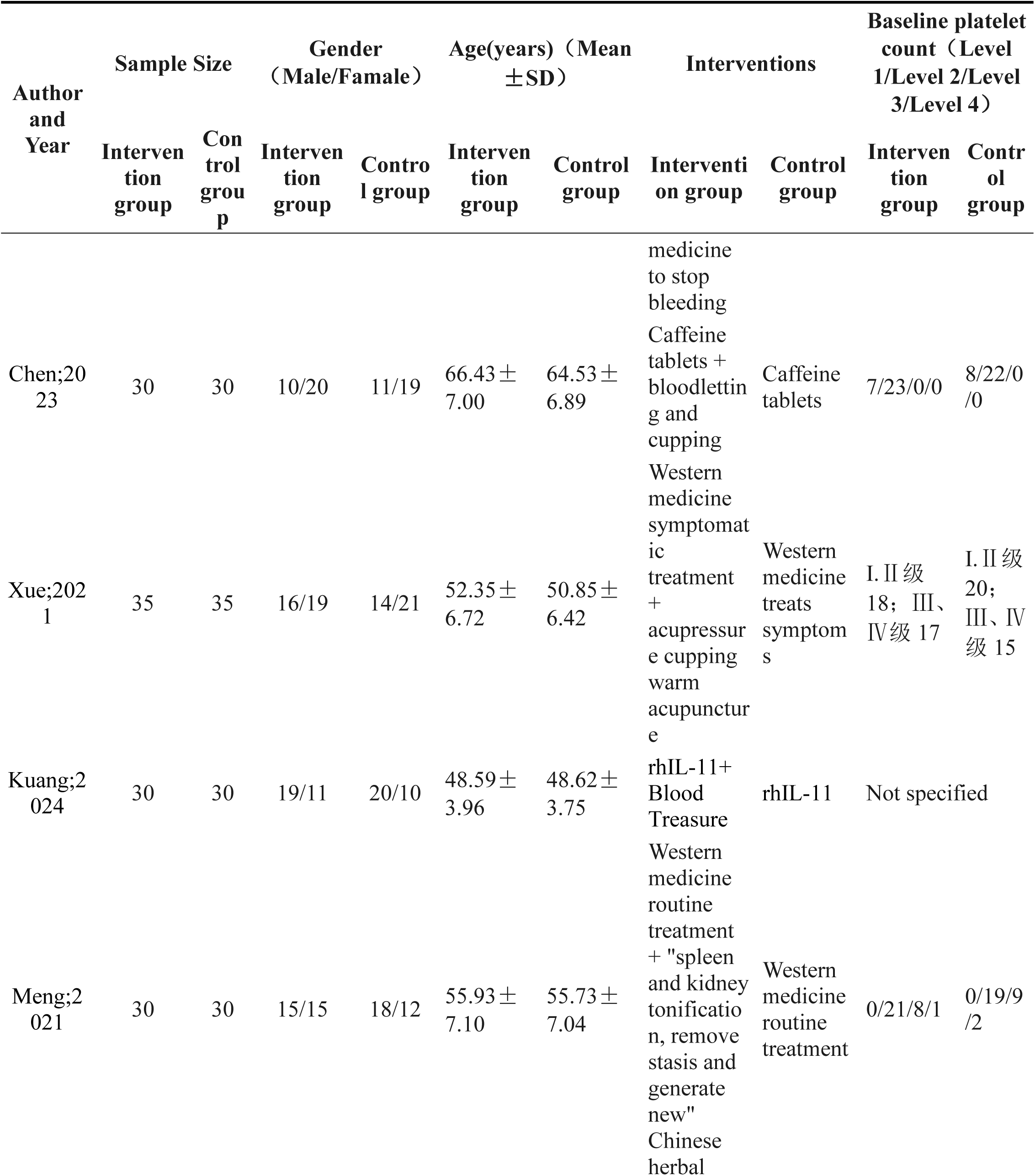
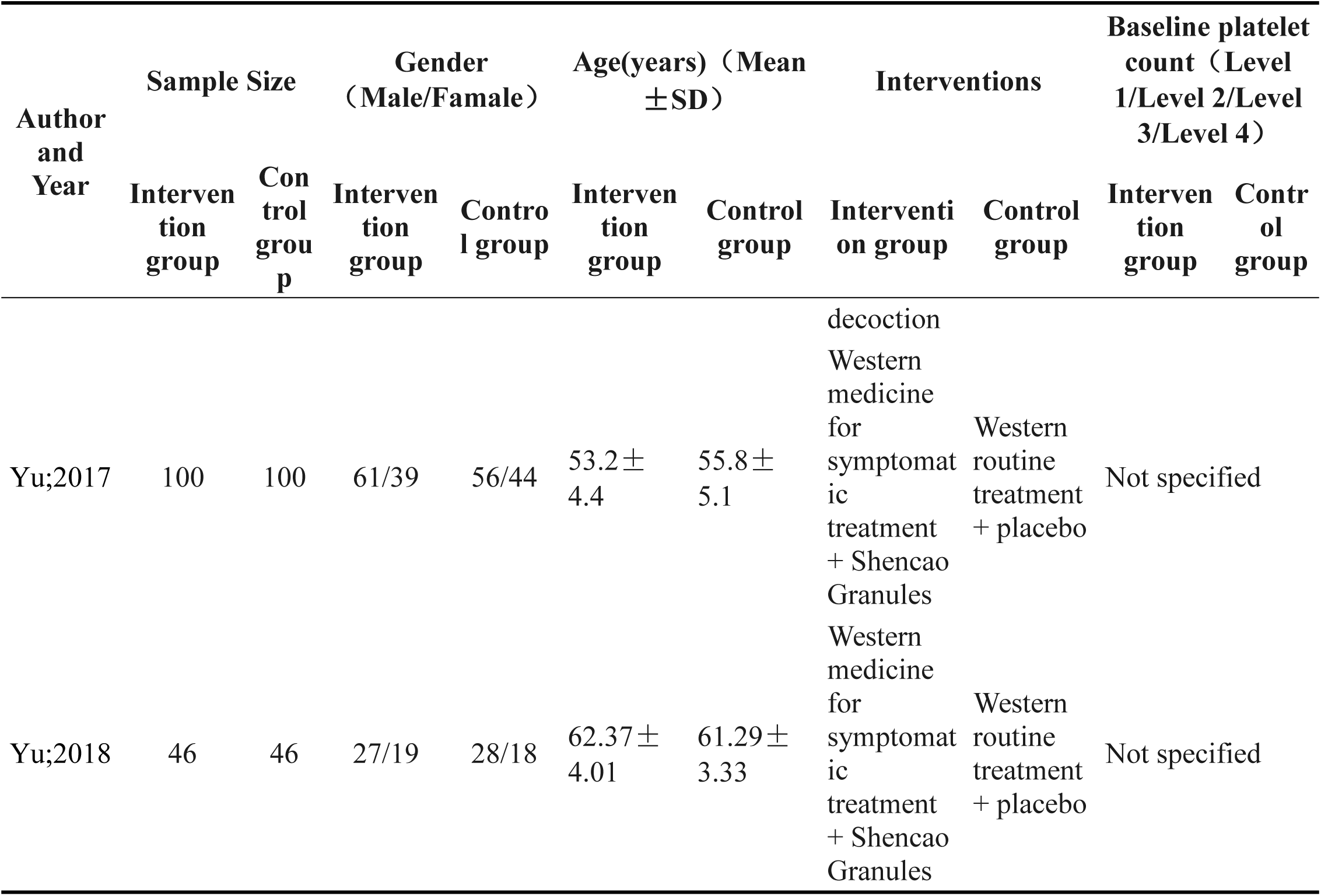
Characteristics of included studies.

### 3.3. Assessment of quality of methodology

All 33 studies reported the use of randomization. Of these, 18 studies did not specify the randomization method, while the remaining studies employed a random digits table. Reporting bias was classified as “unclear,” as the potential risk could not be evaluated due to the unavailability of study protocols for all included trials. None of the 33 studies reported any other sources of bias (Fig. S1).

### 3.4. Clinical outcomes

#### 3.4.1. Significant response rate

Nineteen studies reported a significantly higher response rate for the integrated TCM-WM approach compared to Western medicine alone in the treatment of CTIT. No significant statistical heterogeneity was observed among the included studies (I² = 32.1%, P = 0.089). Consequently, a fixed-effects model was employed for the meta-analysis. The results demonstrated that the experimental group achieved a significantly higher response rate than the control group [RR = 1.54, 95% CI (1.40, 1.69)], as shown in Fig. 2A.A funnel plot was generated for the 19 studies reporting the significant response rate (Fig. 2B). Egger’s test revealed the presence of publication bias (P = 0.000 < 0.05).

**Fig. 2.**
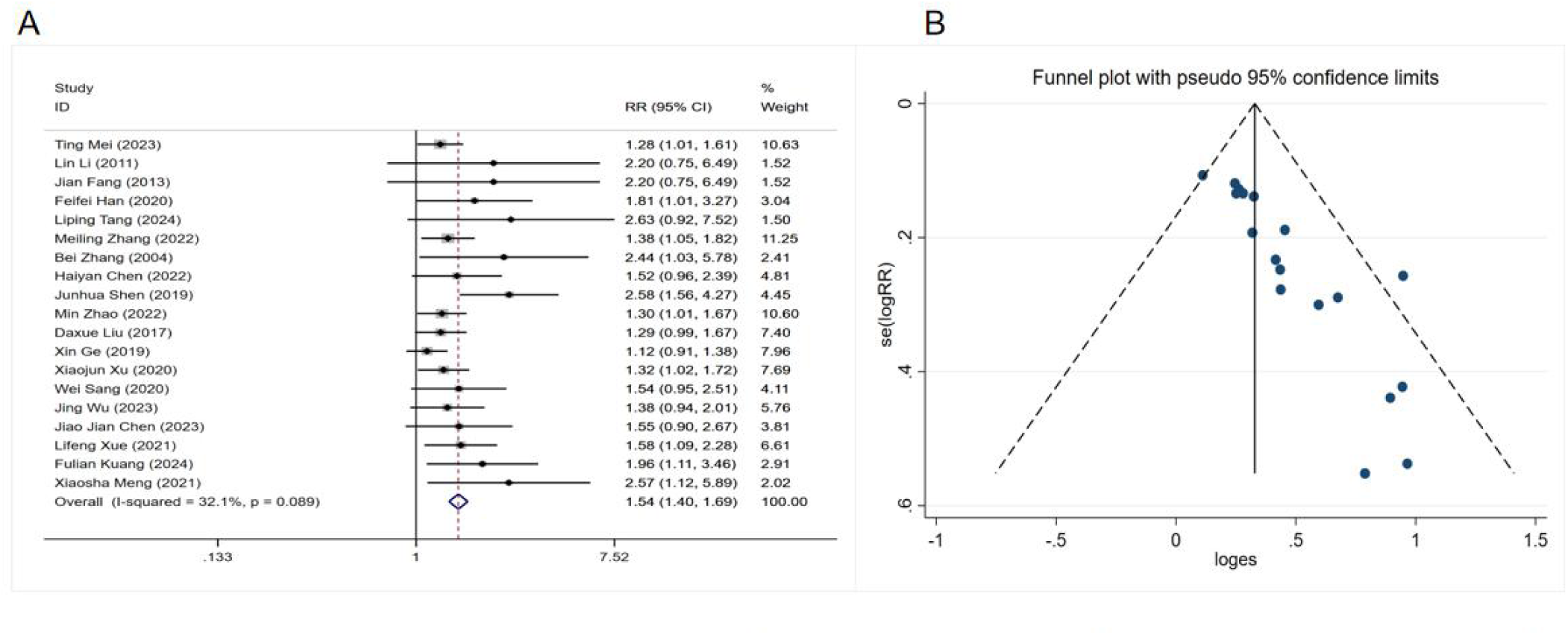
Forest plot (A) and Funnel plot (B) of significant response rate in CTIT patients treated with TCM combined with WM.RR:Relative Risk;CI: confidence interval.

#### 3.4.2. Overall response rate

Twenty studies reported the overall response rate for the integrated TCM-WM approach compared to Western medicine alone in the treatment of CTIT. Significant statistical heterogeneity was observed among the studies (I² = 75.1%, P = 0.000); consequently, a random-effects model was used for the meta-analysis. The results revealed that the experimental group achieved a significantly higher overall response rate than the control group [RR = 1.23, 95% CI (1.14, 1.33), P = 0.000], as shown in Fig. 3A.A funnel plot was generated based on the 20 studies using the overall response rate as the outcome measure (Fig. 3B). It is noteworthy that we also performed a subgroup analysis based on the type of intervention, which revealed that the combination of Traditional Chinese Medicine and TPO-RA resulted in a significantly higher efficacy rate(RR = 1.46, 95% CI: 1.09,1.97)(Fig. 3C).

**Fig. 3.**
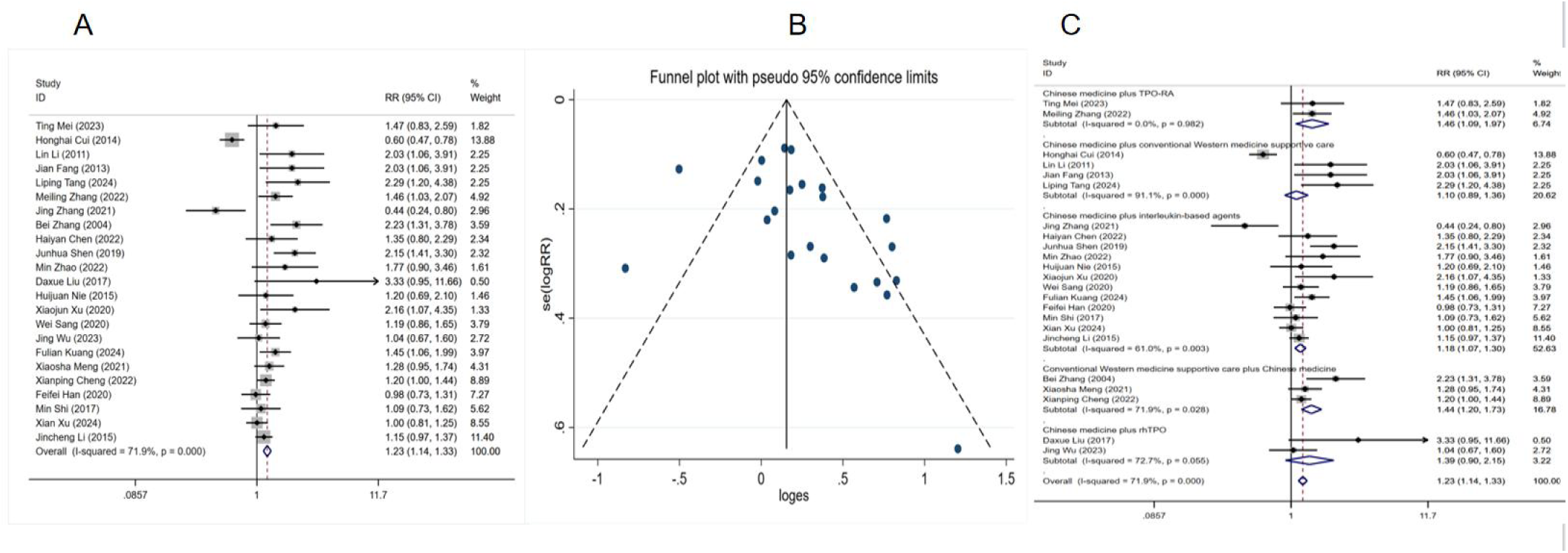
Forest plot(A) „ Funnel plot(B) and Subgroup Analysis(C) of overall response rate in CTIT patients treated with TCM combined with WM.RR:Relative Risk;CI: confidence interval.

Egger’s test suggested the presence of publication bias (P = 0.036 >0.05) (Fig. S2A). To address this, further analysis was performed using the trim-and-fill method. After four iterations, seven studies were estimated to be missing. Following the imputation of these seven hypothetical studies, a repeated meta-analysis was conducted. The heterogeneity test remained significant (P = 0.000), indicating substantial heterogeneity; therefore, a random-effects model was applied for the pooled analysis. The combined odds ratio (OR) was 1.130, consistent with the original conclusion, suggesting that the overall result is robust and not substantially influenced by publication bias.

#### 3.4.3. Duration of thrombocytopenia

Eighteen studies reported the duration of thrombocytopenia following the use of integrated TCM-WM compared to Western Medicine alone for CTIT. Significant statistical heterogeneity was observed among the studies (I² = 92%, P = 0.000), necessitating the application of a random-effects model for the meta-analysis. The results demonstrated that the experimental group experienced a significantly shorter duration of thrombocytopenia compared to the control group [MD = -3.46, 95% CI (−4.30, -2.63)], as shown in Fig. 4A.A funnel plot was generated for the 18 studies, using the duration of thrombocytopenia as the outcome measure (Fig. 4B). The subgroup analysis(Fig. 4C) demonstrated that the 95% confidence intervals of the mean difference for all subgroups were below zero, indicating that each of the three Chinese medicine-integrated regimens significantly outperformed their respective control groups in reducing the duration of thrombocytopenia. In terms of efficacy magnitude, the hierarchy was as follows: conventional Western medicine supportive care combined with Chinese medicine > Chinese medicine plus interleukin-based agents > Chinese medicine plus rhTPO.Egger’s test indicated no evidence of significant publication bias (P = 0.212 > 0.05)(Fig. S2B).

**Fig. 4.**
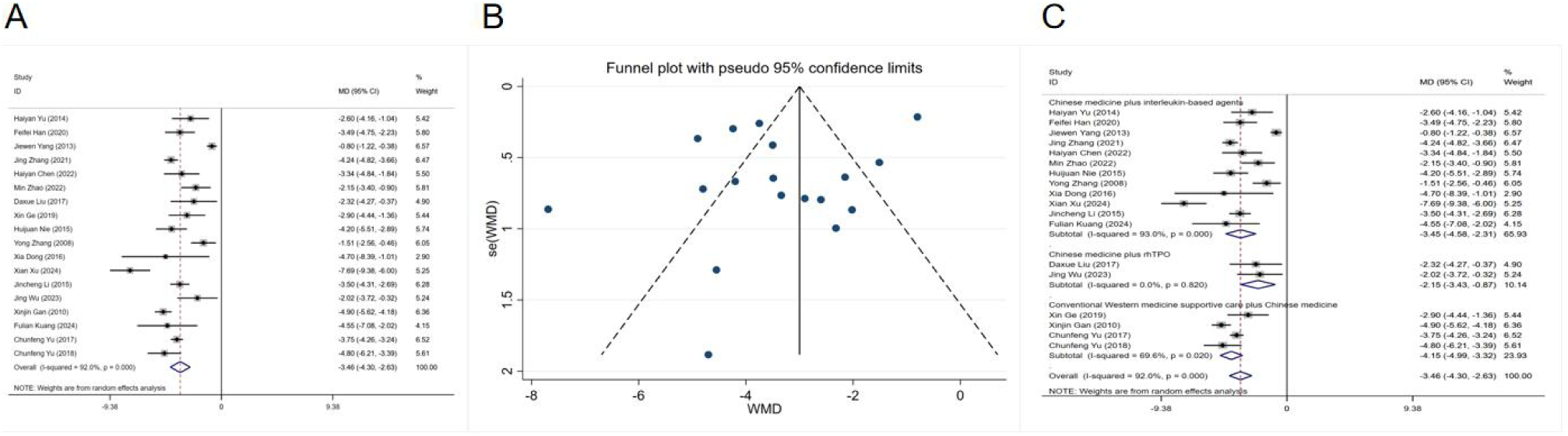
Forest plot(A), Funnel plot(B) and Subgroup Ana!ysis(C) of duration of thrombocytopenia of CTIT patients treated with TCM combined with WM.RR:Relative Risk;CI: confidence interval.

#### 3.4.4. The nadir platelet count

Twenty-one studies reported the nadir platelet count following treatment with integrated TCM-WM compared to Western Medicine alone for CTIT. Considerable statistical heterogeneity was observed across the study results (I² = 86.3%, P = 0.000); therefore, a random-effects model was applied for the meta-analysis. The analysis revealed that the experimental group exhibited a significantly higher nadir platelet count compared to the control group [RR = 13.59, 95% CI (11.01, 16.17), P = 0.000], as shown in Fig. 5A.A funnel plot was generated for the 21 studies, using nadir platelet count as the outcome measure (Fig. 5B). Egger’s test indicated no evidence of significant publication bias (P = 0.021 < 0.05)(Fig. S3A).

**Fig. 5.**
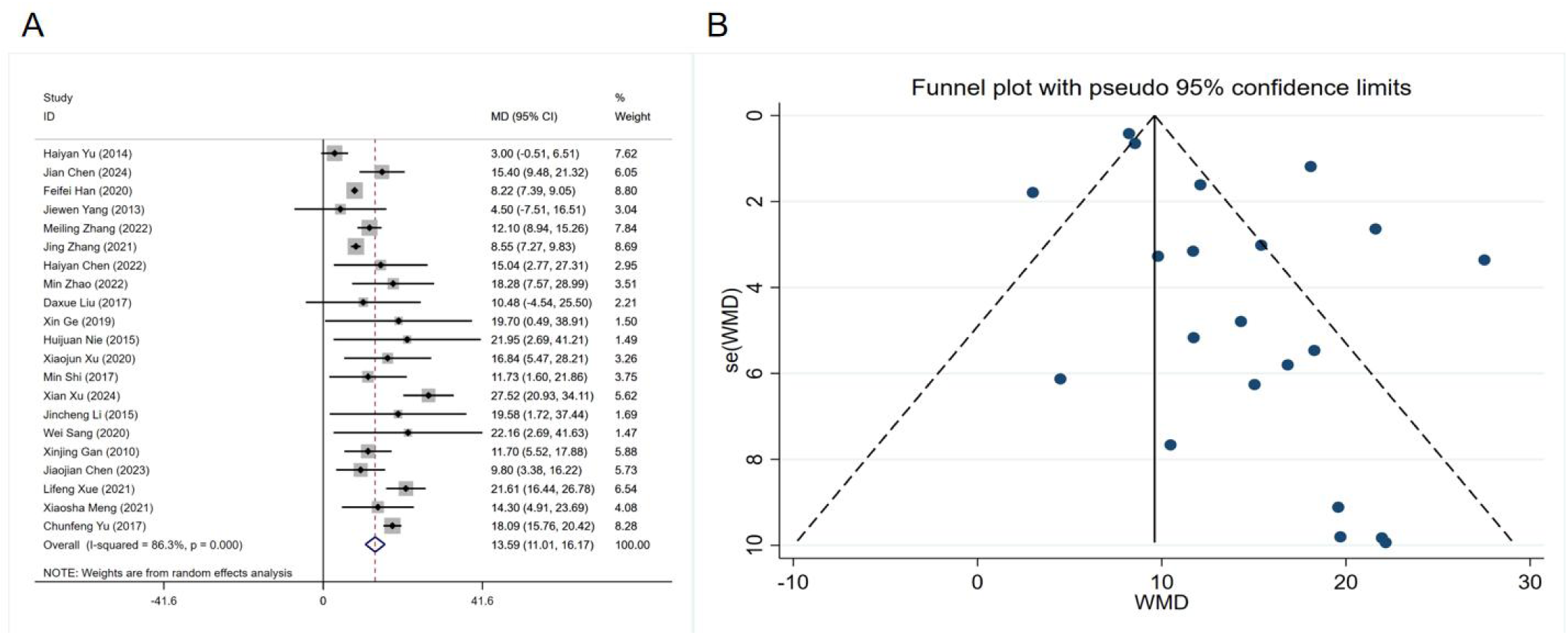
Forest plot(A) and Funnel plot(B) of the nadir piatelet count of CTIT patients treated with TCM combined with WM.RR:Relative Risk;Ci: confidence interval.

#### 3.4.5. Adverse events rate

Four studies reported the incidence of adverse reactions following treatment with integrated TCM-WM compared to Western Medicine alone for CTIT. After excluding the study by Xue Lifeng, the result shifted from non-significant to statistically significant, indicating instability in the meta-analysis outcome. A revised forest plot was generated, demonstrating a higher incidence of adverse events in the experimental group compared to the control group (Fig. 6A). A funnel plot was generated for the 4 studies, using the incidence of adverse events as the outcome measure(Fig. 6B).Egger’s test indicated no evidence of significant publication bias (P = 0.614 > 0.05)(Fig. S3B).

**Fig. 6.**
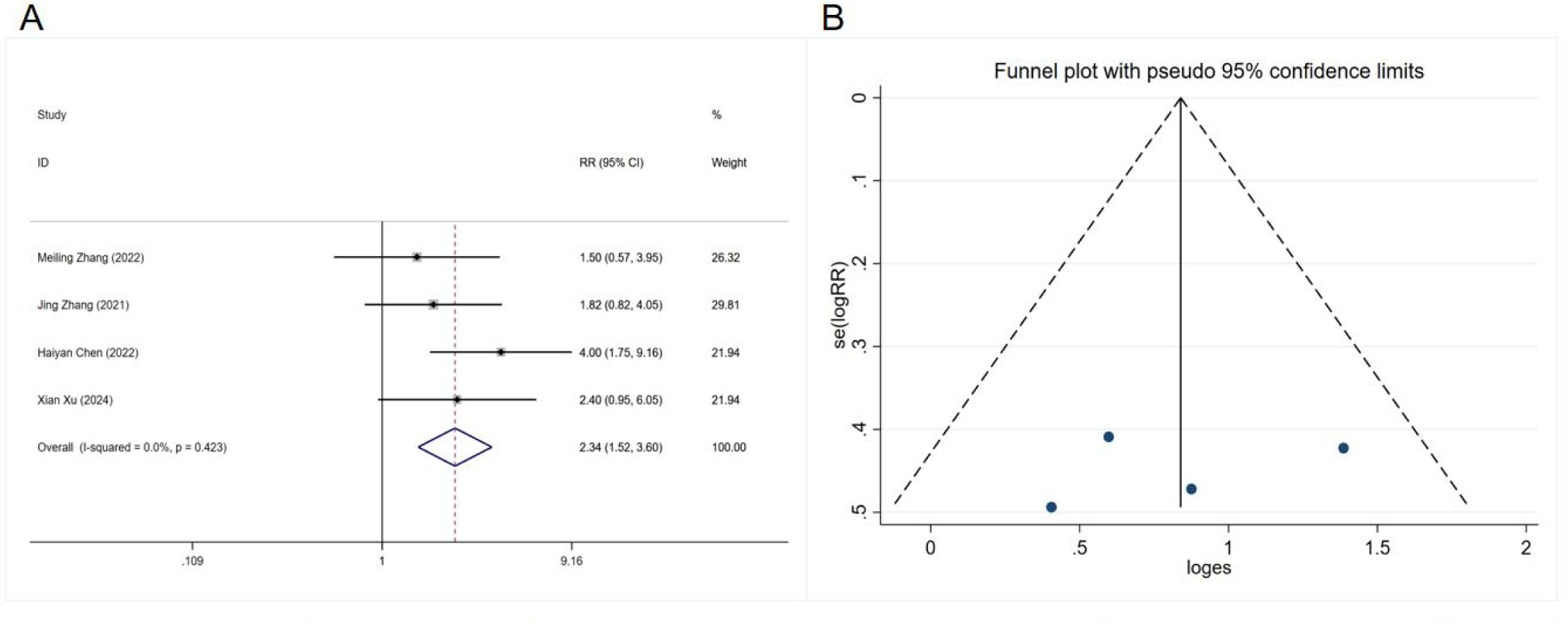
Forest plot(A) and Funnel plot(B) of adverse events rate of CTIT patients treated with TOM combined with WM.RR:Relative Risk;CI: confidence interval.

## 4. Discussion

This meta-analysis highlights the significant advantages of integrating TCM-WM over Western medicine alone in the management of CTIT, as demonstrated across multiple outcome measures, including overall response rate, platelet recovery, duration of thrombocytopenia, and the incidence of adverse events.

From a platelet biology perspective, the underlying mechanisms driving these therapeutic benefits likely involve multiple molecular pathways. Thrombopoiesis is predominantly regulated by thrombopoietin (TPO), a key cytokine that stimulates the proliferation, differentiation, and platelet production of megakaryocytes^[39]^. Numerous chemotherapeutic agents disrupt megakaryopoiesis by damaging hematopoietic stem and progenitor cells in the bone marrow, as well as by impairing the marrow microenvironment essential for platelet production^[40]^. Traditional Chinese Medicine (TCM) interventions may help mitigate these adverse effects through several biological mechanisms.First, specific compounds derived from TCM may enhance the expression of TPO or potentiate the signaling of its receptor, c-MPL. By modulating TPO and its receptor signaling pathway, these compounds may promote the proliferation and differentiation of megakaryocytes, the progenitor cells responsible for platelet production. For instance, studies have demonstrated that compounds such as Radix Astragali and Glycyrrhiza glabra can modulate TPO levels, thus facilitating platelet recovery in thrombocytopenic patients^[41]^.Second, certain herbal ingredients found in TCM formulations may directly stimulate megakaryocyte maturation and promote proplatelet formation. This action could accelerate platelet production, which is often impaired in chemotherapy-treated patients. One such example is Salvia miltiorrhiza, a well-known TCM herb, which has been shown to promote megakaryocyte differentiation in preclinical models, thereby enhancing platelet production. Additionally, compounds like Dang Gui (Angelica sinensis) have been found to support megakaryocyte maturation through the modulation of key signaling pathways that regulate cell differentiation^[42]^.Third, TCM has the potential to protect bone marrow stromal cells, which play an essential role in maintaining the hematopoietic microenvironment. TCM interventions may attenuate the inflammatory damage caused by chemotherapy, which often results in the destruction of the hematopoietic niche and impaired platelet production. For example, Ginseng and Astragalus have demonstrated anti-inflammatory properties that help preserve the integrity of bone marrow stromal cells, reducing chemotherapy-induced damage. By safeguarding these stromal cells, TCM may foster a conducive environment for hematopoiesis, thereby promoting the regeneration of blood cell lineages, including platelets^[43]^. Furthermore, certain TCM formulations may modulate immune function, reducing platelet consumption and extending platelet lifespan^[44]^.

However, several limitations must be acknowledged. First, all included studies were conducted in China, which may limit the generalizability of the findings to other populations. Second, the methodological quality of many trials was suboptimal, with insufficient reporting on randomization procedures, allocation concealment, and blinding, potentially introducing performance and detection biases. Third, substantial heterogeneity was observed across several outcome analyses, likely due to variations in TCM formulations, Western medicine regimens, and patient characteristics. Furthermore, publication bias was identified for certain outcomes, suggesting that smaller studies with negative results may be underrepresented.

Despite these limitations, the application of the trim-and-fill method and sensitivity analyses confirmed the robustness of the primary findings. Future research should focus on rigorously designed, multi-center, double-blind RCTs incorporating standardized TCM interventions and extended follow-up periods. Moreover, biomarker-driven investigations may help identify patient subgroups most likely to benefit from integrated therapy.

## Supporting information

Supplementary data of Systematic Review

## Data Availability

All data produced in the present work are contained in the manuscript

## Acknowledgments

The authors would like to express their sincere gratitude to the researchers whose primary data made this meta-analysis possible. We also acknowledge the support from the Beijing University of Chinese Medicine Library for providing access to the electronic databases.

## Data availability statement

The data that support the findings of this systematic review and meta-analysis are derived from previously published studies, which are cited and listed in the reference section of this article. All extracted data analyzed during this study are included in this published article and its supplementary information files.

