## Supplementary data of Systematic Review for "Efficacy and safety of integrated Chinese and western medicine for cancer therapy-induced thrombocytopenia: A meta-analysis"

**Fig. S1.** Risk of bias summary for all included studies

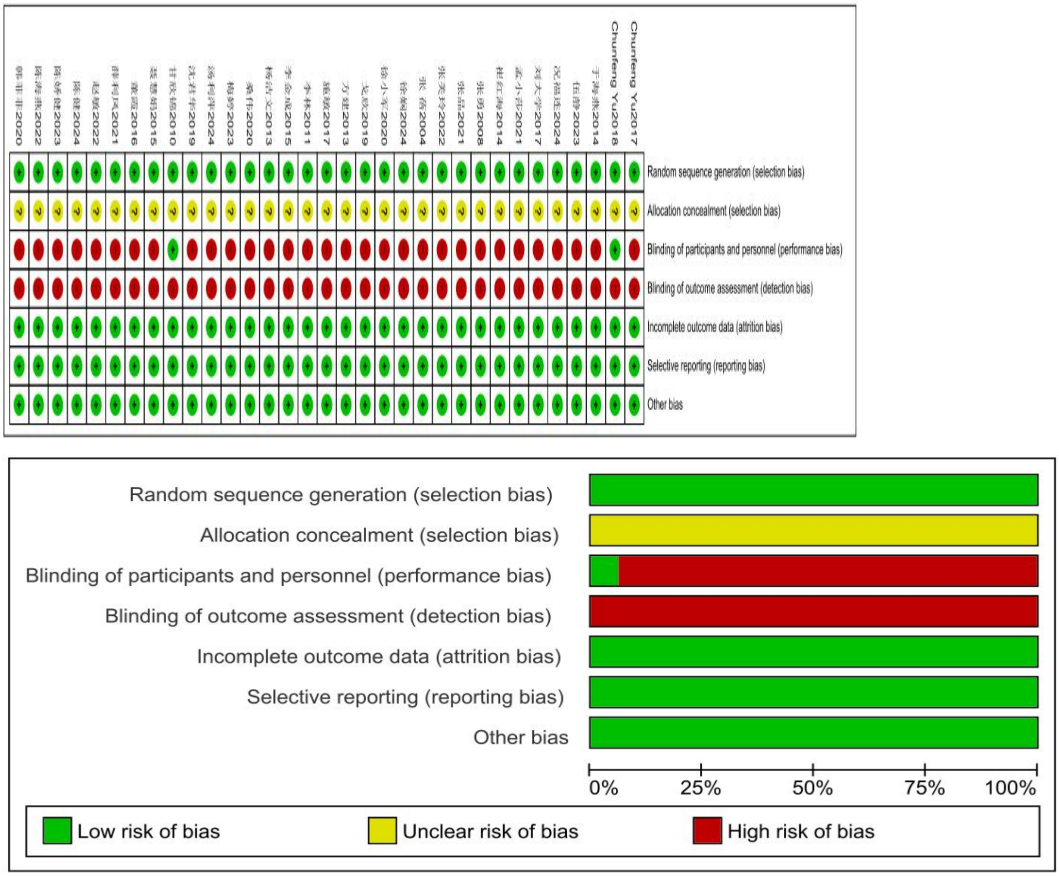

**Fig. S2.** Egger's Test for Publication Bias of the Overall Response Rate and duration of thrombocytopenia.

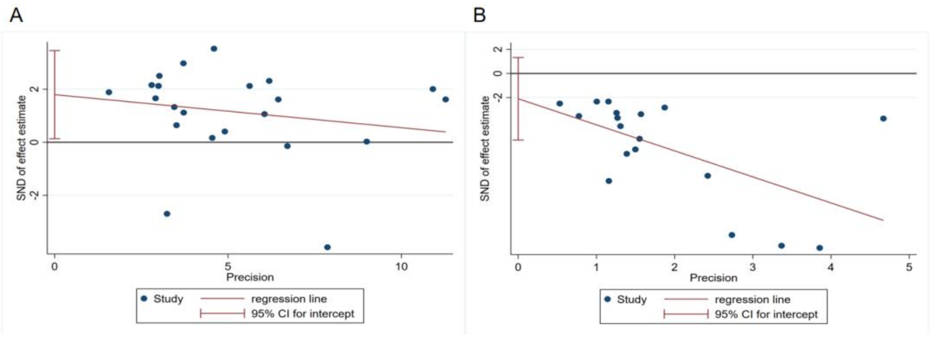

**Fig. S3.** Egger's Test for Publication Bias of the nadir platelet count and adverse events rate.

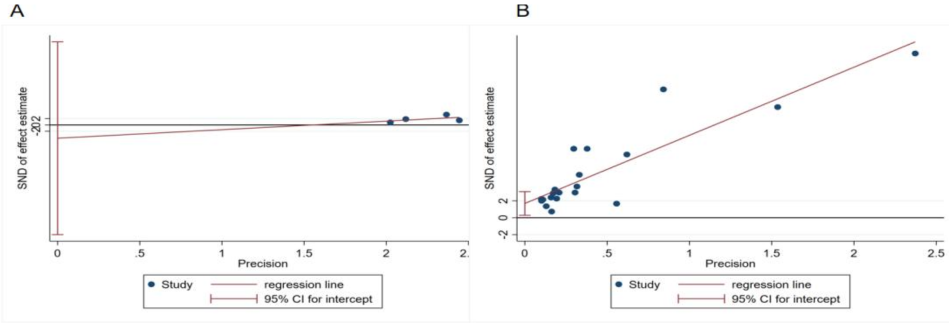

**Table S1.** Search strategy used in each database.

| Database | Search strategy |
| --- | --- |
| PubMed | ((((((("Thrombocytopenia"[Mesh])OR(((Thrombocytopenias[Title/Abstract])OR(Thrombopenia[Title/Abstract]))OR (Thrombopenias[Title/Abstract]))) OR (Chemotherapy induced thrombocytopenia)) OR (immune checkpoint inhibitor induced thrombocytopenia)) OR (targeted therapy-induced thrombocytopenia)) AND (Tumor treatment)) AND (((integrated traditional Chinese and Western medicine) OR (integrated traditional Chinese and Western medicine treatment)) OR (("Medicine, Chinese Traditional"[Mesh]) OR((((((((ZhongYiXue[Title/Abstract])OR(ChungI Hsueh[Title/Abstract])) OR(Hsueh,ChungI[Title/Abstract]))OR(TraditionalMedicine,Chinese[Title/Abstract]))OR(ChineseTraditionalMedicine[Title/Abstract])) OR (Traditional ChineseMedicine[Title/Abstract]))OR(ChineseMedicine,Traditional[Title/Abstract])) OR (Traditional Tongue Diagnosis[Title/Abstract])) OR (Tongue Diagnoses,Traditional[Title/Abstract]))))) AND (((randomized controlled trial[Title/Abstract])OR(randomized[Title/Abstract]))OR(placebo[Title/Abstract])) |
| Cochrane | (Zhong Yi Xue):ti,ab,kw OR (Chung I Hsueh):ti,ab,kw OR (Hsueh, Chung I):ti,ab,kw OR (Traditional Medicine, Chinese):ti,ab,kw OR (Chinese Traditional Medicine):ti,ab,kw OR (Traditional Chinese Medicine):ti,ab,kw OR (Chinese Medicine, Traditional):ti,ab,kw OR (Traditional Tongue Diagnosis):ti,ab,kw OR (Tongue Diagnoses, Traditional):ti,ab,kw OR (Tongue Diagnosis, Traditional):ti,ab,kw OR (Traditional Tongue Diagnoses):ti,ab,kw OR (Traditional Tongue Assessment):ti,ab,kw OR (Tongue Assessment, Traditional):ti,ab,kw OR (Traditional Tongue Assessments):ti,ab,kw |
| Embase | 'Thrombocytopenias':ab,ti or 'Thrombopenia':ab,ti or 'Thrombopenias':ab,ti or 'Chemotherapy induced thrombocytopenia':ab,ti or 'Immune checkpoint inhibitors-induced thrombocytopenia':ab,ti or 'Targeted therapy-induced thrombocytopenia':ab,ti'integrated traditional Chinese and Western medicine treatment':ab,ti or 'traditional Chinese medicine':ab,ti or 'Zhong Yi Xue':ab,ti or 'Chung I Hsueh':ab,ti or 'Hsueh, Chung I':ab,ti or 'Traditional Medicine, Chinese':ab,ti or 'Chinese Traditional Medicine':ab,ti or 'Traditional Chinese Medicine':ab,ti or 'Chinese Medicine, Traditional':ab,ti or 'Traditional Tongue Diagnosis':ab,ti or 'Tongue Diagnoses, Traditional':ab,ti'randomized controlled trial':ab,ti or 'randomized':ab,ti or 'placebo':ab,ti |
| Chinese Biomedical Literature Database | ("random"[Common Fields] OR "randomized controlled"[Common Fields] OR "randomized controlled trial"[Unweighted:Expanded] AND ("integrated traditional Chinese and Western medicine |

| Database | Search strategy |
|---|---|
| | therapy"[Unweighted:Expanded] OR "integrated traditional Chinese and Western medicine"[Unweighted:Expanded] AND ("targeted therapy-related thrombocytopenia"[Common Fields] OR "immunotherapy-related thrombocytopenia"[Common Fields] OR "chemotherapy-induced thrombocytopenia"[Common Fields] OR "cancer therapy-related thrombocytopenia"[Common Fields] OR "thrombocytopenia"[Unweighted:Expanded])) |
| China National Knowledge Infrastructure Database | (Subject: cancer therapy-related thrombocytopenia) OR (Subject: chemotherapy-induced thrombocytopenia) OR (Subject: chemotherapy-related thrombocytopenia) OR (Subject: immunotherapy-related thrombocytopenia) OR (Subject: targeted therapy-related thrombocytopenia) AND (Subject: integrated traditional Chinese and Western medicine) OR (Subject: traditional Chinese medicine) OR (Subject: Chinese herb) OR (Subject: combination therapy) AND (Abstract: randomized controlled (Exact)) OR (Abstract: controlled (Exact)) OR (Abstract: random (Exact)) OR (Abstract: rct (Exact)) |
| China Science and Technology Journal Database | cancer therapy-related thrombocytopenia OR chemotherapy-induced thrombocytopenia OR chemotherapy-related thrombocytopenia + immunotherapy-related thrombocytopenia OR targeted therapy AND integrated traditional Chinese and Western medicine OR traditional Chinese medicine OR Chinese herb AND randomized controlled OR random OR controlled OR random allocation |

**Table S2.** Inclusion criteria and exclusion criteria.

| | | |
|---|---|---|
| Inclusion criteria | Participants | Adults aged 18-75 years, or older, with thrombocytopenia induced by cancer therapy |
| | Interventions | Integration of Traditional Chinese Medicine (TCM) and Western Medicine (WM) therapies |
| | Comparisons | Western Medicine therapies, with or without placebo |
| | Outcomes | The primary outcomes were the significant response rate and overall response rate, as defined by the National Cancer Institute grading criteria for acute and subacute toxicity of anticancer agents |
| | Study design | Randomized controlled trials (RCTs) with two arms, consisting of an intervention group and a control group |
| exclusion criteria | Studies were excluded if they used inappropriate outcome measures for overall response rate or were not published in English or Chinese. | |

**Table S3.** CTIT grading.

| **Level** | **Platelet count (×10^9/L)** |
|---|---|
| 1 | ＜LLN~75 |
| 2 | ＜75~50 |
| 3 | ＜50~25 |
| 4 | ＜25 |
| 5 | platelet reduction-related death |

CTIT:Cancer-therapy-induced thrombocytopenia.LLN:Lower Limit of Normal.
CTIT grading refers to the grading standards for reduced platelet count in the Common Terminology Criteria for Adverse Events (CTCAE) version 5.0 (November 2017).
